# Post-Discharge Anti-Seizure Medication Use Improves Post-Stroke Survival: *An Emulated Target Trial in Older Adults*

**DOI:** 10.64898/2026.04.17.26351149

**Authors:** Madhav Sankaranarayanan, Maria A. Donahue, Julianne D. Brooks, Shuo Sun, Joseph P. Newhouse, Deborah Blacker, Sebastien Haneuse, Sonia Hernández Díaz, Lidia M.V.R. Moura

## Abstract

**Objective:** Levetiracetam is commonly prescribed for seizure prophylaxis after acute ischemic stroke (AIS) and often continued beyond discharge. While its short-term effectiveness for preventing post-stroke seizures is established, it is unclear whether prolonged use improves survival, particularly in older adults. We estimated the effect of continued levetiracetam use on 90-day mortality among Medicare beneficiaries after AIS.

**Methods:** Using Traditional Medicare claims data (2008-2021), we identified beneficiaries aged ≥66 years hospitalized for AIS who initiated outpatient levetiracetam within 90 days of discharge. After one month of continued post-stroke use of levetiracetam (start of follow-up), we compared 90-day mortality between patients with a new levetiracetam dispensation within a 14-day grace period post-follow up and those without one. We performed cloning, censoring and weighting to address immortal time bias and estimated standardized mortality risks, risk differences, and 95% confidence intervals (CI).

**Results:** Among 3,212 eligible beneficiaries, 1,779 (55.4%) received a new levetiracetam dispensation within the 14-day grace period. Median age was 76 years (IQR 70-83); 57.8% were female. After adjustment for demographics, hospitalization characteristics, timing of initiation, and comorbidities, continued use was associated with lower 90–day mortality than discontinuation (53 vs 62 deaths per 1,000; risk difference −9 per 1,000; 95% CI: (−12,−5)). The reduction was observed primarily among patients aged ≥75 years.

**Significance:** Among older Medicare beneficiaries who initiated levetiracetam after AIS, continued outpatient use was associated with modestly lower 90–day mortality, particularly in those aged ≥75 years. These findings suggest potential benefits of levetiracetam continuation beyond the immediate post-stroke period.

## INTRODUCTION

Poststroke seizures are a known complication of acute ischemic stroke (AIS), particularly in older adults.^1^ For AIS survivors who are prescribed an antiseizure medication (ASM), levetiracetam is the most common, and roughly 40% of older adults who initiate levetiracetam within 30 days of AIS discharge remain adherent over the subsequent year, with the remainder discontinuing or becoming non–adherent within months.^2–4^ In real-world practice, a substantial proportion of older adults initiated on levetiracetam after AIS remain on therapy for prolonged periods, often in the absence of a clearly documented long-term indication.^5^

Levetiracetam has become the dominant ASM in poststroke care due to its favorable pharmacokinetic profile, minimal drug-drug interactions, and relative ease of use in older adults.^3^ Once levetiracetam is started during the poststroke period, it is often continued by default, resulting in treatment persistence among older adults.^6,7^ However, it is not without clinically meaningful adverse effects, including mood disturbances, irritability, fatigue, gait instability, and cognitive changes, outcomes that are particularly salient in a vulnerable poststroke population but are not reliably captured in administrative data.^8^

The clinical rationale for continued ASM use after stroke rests on the assumption that ongoing treatment reduces the risk of seizures and, by extension, life-threatening complications, including mortality.^9,10^ However, there is a lack of robust evidence to support the claim that continued use of levetiracetam after hospital discharge improves survival among poststroke patients.^6,11^

In this study, we used an emulated target trial framework applied to a large, nationally representative sample of Medicare beneficiaries to estimate the effect of continued outpatient levetiracetam use on 90-day mortality among older adults who were prescribed levetiracetam after AIS discharge. Beyond the specific clinical question, this study aims to illustrate how emulated target trials can generate valid evidence for clinically consequential questions that cannot be ethically or feasibly addressed through randomized trials in vulnerable populations.

## METHODS

### Study Design

We used a target-trial approach to emulate a hypothetical randomized clinical trial.^12,13^ We defined an ideal target trial (**Table 1**) to provide a conceptual framework for estimating the effect of outpatient prescription continuation. We will be considering patients who have had a prespecified period of continued levetiracetam usage, and we define the time zero as the end of this period. The hypothetical target trial randomly assigned eligible patients to one of two arms: i) prescription of levetiracetam at time zero for one month (which we refer to as the continuation arm), and ii) no use of levetiracetam for one month after time zero (which we refer to as the discontinuation arm). We provide a framework for target trial emulation for this hypothetical trial by carefully defining our study population using our data sources (see **Table 1** for more details on the emulation).

**Table 1.** Transparent Reporting of Observational Studies Emulating a Target Trial (The TARGET Statement)

| Target Trial | Emulated trial |
| --- | --- |
| **Eligibility Criteria** |  |
| Discharge for cerebrovascular accident* between 04/01/2008 and 09/30/2021 from any hospital, and enrolled in Traditional Medicare Part A, B, D 12 months before stroke admission. | Same |
| Age $\geq 66$ at discharge | Same |
| Discharged from a short-stay hospital | Same |
| No previous history of AIS in the 12 months before admission. | No recorded diagnosis of AIS in the last 12 months before admission |
| No use of levetiracetam in the 3 months prior to admission. | No recorded prescription of levetiracetam in the last 3 months before admission. |
| Prescribed levetiracetam at discharge and had continued usage for an initial usage window of 1 month. | Prescription for levetiracetam within 90 days post discharge from AIS admission.<br>Prescription fills cover 50% of days from the initial usage window of 1 month.<br>Refill gaps between prescriptions below 14 days in the initial usage window of 1 month.<br>Last dispensation within the initial usage window has a supply that ends within 7 days before or 30 days after the end of the initial usage window of 1 month. |
| **Treatment Strategies** |  |
| Time zero: End of initial usage window | Time zero: End of initial usage period |
| Treatment arm: New outpatient prescription of levetiracetam to be filled within a 14-day window following time zero | Treatment arm: At least one outpatient prescription of levetiracetam within a 14-day continuation grace period following time zero. |
| Control arm: No outpatient prescription of levetiracetam after time zero. | Control arm: No outpatient prescriptions of levetiracetam within 14 days after time zero. |
| **Assignment Procedures** |  |
| Randomized treatment assignment | Emulated randomization using cloning* |
| **Outcomes** |  |
| Time to death within 90 days from time zero | Same |
| **Follow-up** |  |
| Starts at randomization (time zero) | Starts at time zero (end of initial usage period) |
| Ends at date of death, loss to follow-up or end of study (90 days after time zero), whichever occurs first. | Ends at date of death (as recorded in Medicare data<sup>†</sup>), loss to follow-up (all-cause readmission<sup>‡</sup>), or end of study (90 days post-discharge), whichever occurs first. |
| **Causal Contrast** |  |
| Per-protocol effect | Observational analog of per-protocol effect<sup>§</sup> |
| **Identifying Assumptions and Statistical Analysis** |  |
| Random treatment assignment, no unmeasured confounding, and correct model specification | Conditional exchangeability after adjustment, no unmeasured confounding, and correct model specification |
| Per-protocol effect analysis of time to mortality,<br>accounting for censoring. | Per-protocol effect analysis of time to mortality, accounting<br>for censoring and baseline confounding. |

**Legend:** AIS, Acute Ischemic Stroke; \*Further details are provided in Supplement. <sup>†</sup>Medicare data mortality: Dates obtained from the Master Beneficiary Summary File, a reliable metric for mortality. <sup>‡</sup>Readmissions are a loss to follow-up; we do not have insurance claims information for inpatients, and dates were obtained from the Medicare Provider Analysis and Review file. <sup>§</sup>Analysis can be consistent with “per-protocol” (when treatment strategy is considered a “protocol” that must be followed over time [grace period]), as we cannot get “intention-to-treat” (treatment assignment as a strategy).

### Setting & Data Sources

We used insurance claims data from the Medicare Master Beneficiary Summary (MBSF) File.^14^ The file contains information on demographic characteristics, including age, sex, race, and Medicare and Medicaid eligibility status. We also used acute hospitalization records from the Medicare Provider Analysis and Review (MEDPAR)^15^ file and outpatient diagnosis codes in the Outpatient and Carrier Claims files. The MEDPAR files contain information on hospitalization dates and reasons for admission, while the Outpatient and Carrier Claims files contain information on prescription fills. Readmission data is provided in the MEDPAR file, and death dates are in the MBSF. This study followed the Strengthening the Reporting of Observational Studies in Epidemiology (STROBE; Table S5)^16^ and the Transparent Reporting of Observational Studies Emulating a Target trial (TARGET)^17^ reporting guidelines (Table 1).

### Eligibility Criteria

In a 20% random sample of Traditional Medicare Claims, we identified 224,991 patients who were admitted for AIS between April 1, 2008, and September 30, 2021, and with at least 12 months of enrollment in the Traditional Fee-For-Service Medicare Parts A (hospital insurance information), B (preventative and medically necessary services or medical insurance information), and D (prescription drug coverage information) before AIS hospitalization. We describe our inclusion criteria in Figure 1. We included patients aged 66 and above who were admitted to a hospital and discharged home (short-stay hospitalization). We excluded patients with a recorded AIS diagnosis within 12 months of hospitalization and those with one or more recorded outpatient dispensations for levetiracetam within three months of admission.

**Figure 1.**
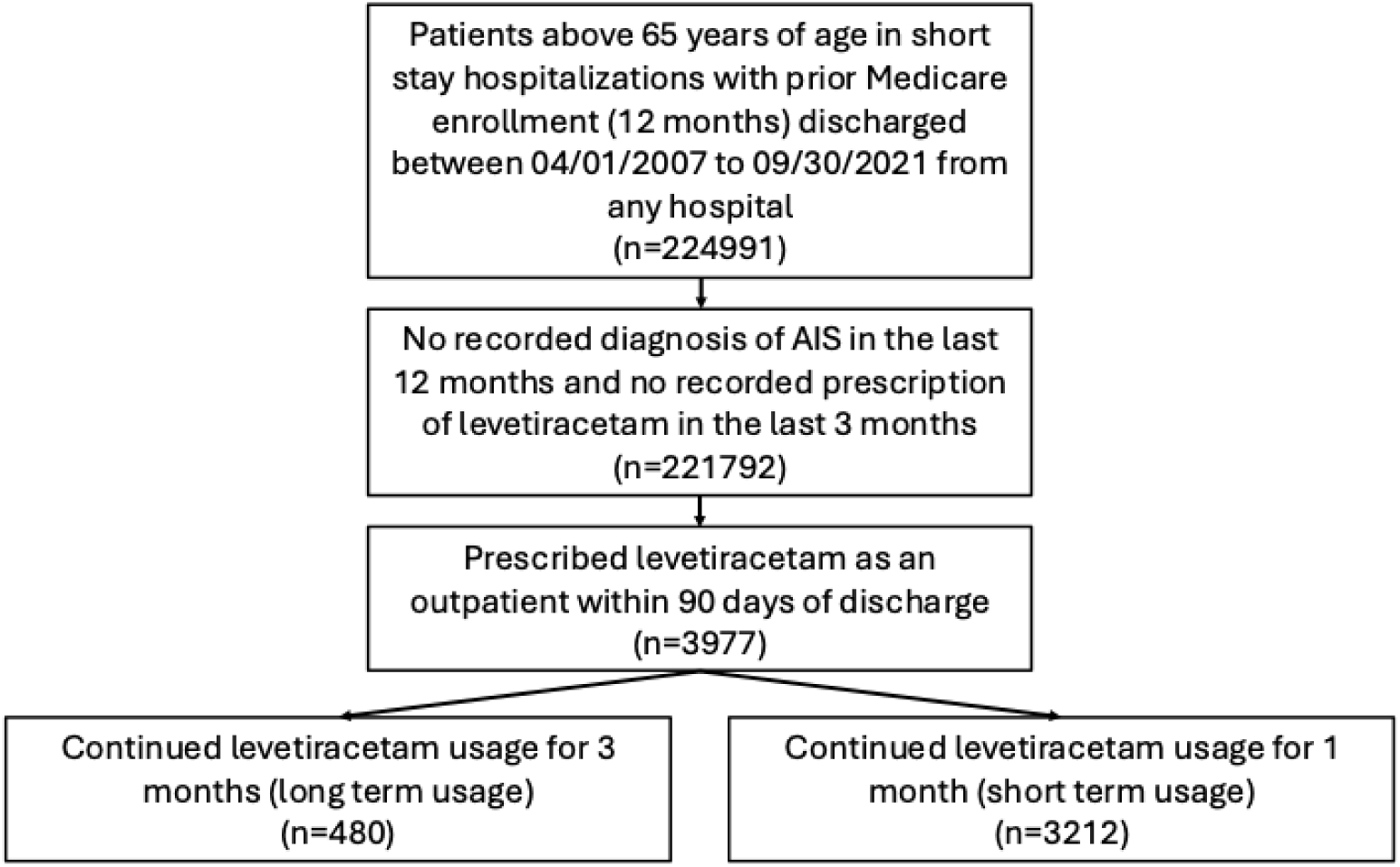
Schematic Describing Eligibility Criteria. **Legend:** Describes the sampling process resulting in samples of 480 and 3212 patients. Medicare data files used: MedPAR (Inpatient data), MBSF (Summary data); Diagnosed for Acute Ischemic Stroke based on ICD-10 codes (I63 and I63.9); Part A: Hospital Insurance, Part B: Medical Insurance; Part D: Drug coverage. AIS, Acute Ischemic Stroke; FFS, Fee-for-Service.

We further restricted the population to those who received at least one dispensation for levetiracetam within 90 days of discharge from the AIS admission. We identified levetiracetam dispensations from outpatient pharmacy data for the target trial emulation. For eligibility, when prescriptions overlap, we stacked them under the assumption that patients retain any excess medication beyond the end of each prescription. We required continued outpatient usage during an initial usage window for our eligible sample. We defined the initial usage window as the period after the first post-discharge levetiracetam dispensation, during which we applied the inclusion criteria based on initial usage. For our main analysis, this initial usage window is one month (**Figure S1**). We defined the time covered by dispensations within this initial usage window as the initial usage period, noting that it varies across patients. For example, in **Figure S1**, the patient has a dispensation that starts within the initial usage window but ends after the window. The end of this dispensation is taken as time zero for this patient. We defined the end of days supplied (end of initial usage period) as our time zero. The following conditions describe continued outpatient usage: (1) prescription fills cover 50% of days in the initial usage window, (2) refill gaps between prescription fills are under 14 days within the initial usage window, and (3) the last dispensation within the initial usage window has a supply that ends within 7 days before or 30 days after the end of the initial usage window. We describe these inclusion criteria in **Figure S1** in the Supplement. We have an eligible sample of 3,212 patients.

We also use a 3-month initial usage window to obtain a sample of 480 patients. We refer to these samples as the short-term and long-term initial usage samples, respectively. We will focus the remaining analyses on the short-term initial usage sample and relegate the study of the long-term initial usage to the “Additional Analyses” in the Supplement.

### Treatment Strategies

To operationalize the target trial in the real-world data, we utilized outpatient dispensations to account for prescriptions. To do so, we use a 14-day grace period from time zero, to allow for patients to refill prescriptions. With this, we identified the treatment arms as: i) at least one outpatient dispensation of levetiracetam of at least a one-month supply within the 14-day grace period after time zero; ii) no outpatient prescriptions of levetiracetam within the 14-day grace period after time zero.

We treated readmission as an administrative censoring mechanism for both arms. We addressed immortal time bias using cloning techniques.^12,13,18^ Briefly, for each patient, a “clone” is created, with one from each pair “assigned” to one of the two treatment arms; key to this step is that the two treatment arms are completely balanced with respect to baseline covariates. Person-time during which outcomes are ascertained is then artificially censored at the first time either clone deviates from the treatment corresponding to their arm. For example, if in the original observation for a given patient was that they did not receive a new outpatient prescription of levetiracetam within the 14-day grace period, their “clone” (who was assigned to the arm in which an outpatient prescription of levetiracetam within 14 days was received) would have their person time censored at day 14. In subsequent analyses, we accounted for the bias induced by this artificial censoring using stabilized inverse probability of censoring weighting (IPCW; **Figure S2** and “Technical Details” in the Supplement).

### Emulated Randomization & Covariates

In the target trial, we would be able to balance baseline characteristics through randomization. For the emulation procedure, we ascertained the distributions of baseline characteristics in the treatment and control populations and adjusted for potential confounders. We describe the individual characteristics of our samples. For each beneficiary, we extracted demographic characteristics, including age, documented sex, reported race/ethnicity, dual Medicare and Medicaid eligibility status, and the original reason for Medicare entitlement as reported in Medicare’s Master Beneficiary Summary (MBSF) File. We used the self-reported race/ethnicity variable provided in MBSF (White, Black, Hispanic, Asian, American Native, Other, Unknown).^19^

We used acute hospitalization records from the MedPAR file and outpatient diagnosis codes in the Outpatient and Carrier Claims files. AIS hospitalizations were selected based on the AIS ICD-10 code I63, “Cerebral Infarction.”^20^ We took the first hospitalization chronologically for beneficiaries with more than one AIS hospitalization within our study window. We defined readmission as any inpatient stay recorded in the MedPAR file after the index admission. We also obtained baseline stroke severity using the modified Rankin Scale (mRS).^21^ The mRS has seven categories: 0 (no symptoms), 1 (no significant disability), 2 (slight disability), 3 (moderate disability), 4 (mild to severe disability), 5 (severe disability), 6 (death).^22^ This study employed a validated claims-based algorithm to classify mRS as a binary outcome, grouping scores 0-3 as minor disability and 4-6 as moderate to severe disability.^23^ Prior work on the same population has shown that the algorithm accurately identifies disability status with an ROC AUC of 0.85.^23^ As an additional proxy for baseline stroke severity, we used hospital length of stay (LOS) during the index AIS admission, as prolonged LOS in AIS has been associated with worse functional outcomes and a higher risk of adverse events.^24,25^

We analyzed all comorbid conditions present in the 12 months before stroke admission. We evaluated for each beneficiary the presence of a pre-admission history of myocardial infarction (MI), chronic kidney disease (CKD), congestive heart failure (CHF), peripheral vascular disease (PVD), cardiovascular disease (CVD), dementia, and other comorbidities^26^ using ICD-10 diagnosis codes listed in Supplemental Table S1 and the MBSF 30 CCW Chronic Conditions documentation.^39^ This algorithm required that, for outpatient claims, a patient’s diagnoses must appear on at least two claims (codes need not be identical but consistent), with at least 30 days between them. The comorbidities were also combined using the Charlson Comorbidity Index (CCI).^1^

### Follow-up & Outcome: 90-day Mortality

We assessed the primary outcome, mortality, over a 90-day follow-up period from time zero. Patients were followed from time zero to the end of study (i.e., 90 days from time zero, including day zero) or mortality, whichever came first. Ninety days is a widely accepted standard time frame in acute stroke and cardiovascular outcome research, capturing the majority of deaths attributable to the index event and its immediate complications while limiting loss to follow–up and competing long–term causes.^27–29^ Further details on the proposed target trial and the emulation with observational data are provided in **Table 1**. All-cause readmission served as a censoring mechanism during the 14-day grace period; however, it was not considered a loss to follow-up for the remainder of the observation period (since mortality was the primary outcome and can be ascertained from other sources).

### Statistical Analysis

We estimated patients’ survival probabilities using model-based predictions to evaluate the effect of levetiracetam continuation during the defined exposure period on 90-day mortality. The models for IPCW were pooled linear logistic regressions over person-days, including as covariates length of the initial usage period, time to first dispensation post-discharge, age, race/ethnicity, CCI, and LOS, fit separately for the two treatment strategies. We also estimated unadjusted Kaplan-Meier and spline-based survival probabilities of mortality over 90 days using the cloned dataset (**Figure S4** in the Supplement). We also ran analyses with pooled linear logistic regressions with different time scales and a Cox proportional hazards model to examine the robustness of our findings (Additional Analyses in the Supplement).

We fitted a pooled logistic regression model for mortality as a function of the following covariates: treatment strategy, time (days after time zero), and an interaction term between treatment strategy and time (days after time zero) to allow for time-varying effects, using the expanded weighted data. We allowed for flexible specification of the time variable’s function and chose a baseline hazard function by minimizing Akaike Information Criterion (AIC; **Table S4**). The specific functions for the main analysis and each secondary analysis are provided in the Supplementary Materials. We obtained estimated survival probabilities for each day under each treatment strategy and estimated the standardized 90-day mortality risk difference (RD). We used the bootstrap method with 500 resamplings to obtain 95% confidence intervals (CIs), accounting for cloning-based sample-size inflation (Technical Details in the Supplement).

### Missing Data & Additional Pre-Planned Analyses

In accordance with methodological and regulatory guidance, we conducted subgroup analyses by age (a key prognostic factor) to assess potential age–related heterogeneity in the association between levetiracetam continuation and mortality.^30^ Our samples had no missingness for age, sex, race, baseline covariates, and mortality information. We repeated the analyses above on the long-term initial usage sample. We stratified the short-term sample by age categories (Supplement **Tables S2 and S3**, **Figures S5 and S6**, and the Technical Details section). We analyzed the long-term initial-use sample to understand the effects of different continuation patterns.

## RESULTS

### Study Population Characteristics

We examined a sample of 3,212 beneficiaries discharged for an AIS during the study period (**Figure 1**). 1779 (55.4%) patients had a new dispensation for levetiracetam within the 14-day grace period. There were 130 (4.0%) patients who passed away within the 90-day observation period. The median age of our study sample was 76 (IQR: 70-83). We described the characteristics of eligible patients in the short-term initial-use sample, stratified by treatment strategy, in **Table 2**.

**Table 2.** Characteristics of Patients Stratified by Treatment Strategy in the Short-Term Initial Usage Sample.

| Characteristics | Overall Sample | Levetiracetam Continuation |  | SMD |
| --- | --- | --- | --- | --- |
|  |  | No (n=1433) | Yes (n=1779) |  |
| Time to First Dispensation (median (IQR)) | 13 (34) | 17 (56) | 10 (24) | 0.540 |
| Length of Initial Usage Period (median (IQR)) | 29 (1) | 29 (1) | 29 (1) | 0.070 |
| Age (median (IQR)) | 76 (13) | 76 (14) | 76 (13) | 0.061 |
| Age Categories (%) |  |  |  |  |
| 66-74 | 860 (42.1) | 127 (44.3) | 733 (41.8) | 0.146 |
| 75-84 | 776 (38.0) | 93 (32.4) | 683 (38.9) |  |
| 85+ | 405 (19.8) | 67 (23.3) | 338 (19.3) |  |
| Sex (%) |  |  |  |  |
| Male | 1361 (42.4) | 606 (42.3) | 755 (42.4) | 0.003 |
| Female | 1851 (57.6) | 827 (57.7) | 1024 (57.6) |  |
| Race/Ethnicity (%) |  |  |  |  |
| White | 2504 (78.0) | 1092 (76.2) | 1412 (79.4) | 0.127 |
| Black | 485 (15.0) | 239 (16.7) | 244 (13.7) |  |
| Asian | 53 (1.7) | 19 (1.3) | 34 (1.9) |  |
| Hispanic | 94 (2.9) | 49 (3.4) | 45 (2.5) |  |
| American Native | <11 (*) | <11 (*) | <11 (*) |  |
| Other | 40 (1.2) | 20 (1.4) | 20 (1.1) |  |
| Unknown | >27 (*) | >11 (0.9) | >13 (1.0) |  |
| US Regions (%) |  |  |  |  |
| Midwest | 691 (23.4) | 290 (22.0) | 401 (24.6) | 0.11 |
| Northeast | 665 (22.5) | 284 (21.5) | 381 (23.3) |  |
| Southeast | 851 (28.8) | 380 (28.8) | 471 (28.9) |  |
| Southwest | 331 (11.2) | 164 (12.4) | 167 (10.2) |  |
| West | 412 (14.0) | 200 (15.2) | 212 (13.0) |  |
| Year of Discharge (%) |  |  |  |  |
| 2007 | 67 (3.3) | <11 (2.1) | >56 (3.5) | 0.492 |
| 2008 | 91 (4.5) | <11 (3.1) | >80 (4.7) |  |
| 2009 | 96 (4.7) | <11 (3.5) | >85 (4.9) |  |
| 2010 | 102 (5.0) | 11 (3.8) | 91 (5.2) |  |
| 2011 | 139 (6.8) | 14 (4.9) | 125 (7.1) |  |
| 2012 | 126 (6.2) | <11 (2.4) | >115 (6.8) |  |
| 2013 | 146 (7.2) | 21 (7.3) | 125 (7.1) |  |
| 2014 | 148 (7.3) | 18 (6.3) | 130 (7.4) |  |
| 2015 | 166 (8.1) | 15 (5.2) | 151 (8.6) |  |
| 2016 | 160 (7.8) | 22 (7.7) | 138 (7.9) |  |
| 2017 | 167 (8.2) | 16 (5.6) | 151 (8.6) |  |
| 2018 | 167 (8.2) | 27 (9.4) | 140 (8.0) |  |
| 2019 | 170 (8.3) | 39 (13.6) | 131 (7.5) |  |
| 2020 | 152 (7.4) | 31 (10.8) | 121 (6.9) |  |
| 2021 | 144 (7.1) | 41 (14.3) | 103 (5.9) |  |
| Baseline Dementia (%) | 785 (24.4) | 389 (27.1) | 396 (22.3) | 0.113 |
| History of MI (%) | 293 (9.1) | 139 (9.7) | 154 (8.7) | 0.036 |
| Atrial Fibrillation (%) | 875 (27.2) | 376 (26.2) | 499 (28.0) | 0.041 |
| Cataract (%) | 2227 (69.3) | 1010 (70.5) | 1217 (68.4) | 0.045 |
| CKD (%) | 1494 (46.5) | 695 (48.5) | 799 (44.9) | 0.072 |
| COPD (%) | 1139 (35.5) | 533 (37.2) | 606 (34.1) | 0.065 |
| CHF (%) | 1362 (42.4) | 640 (44.7) | 722 (40.6) | 0.083 |
| CVD (%) | 2148 (66.9) | 989 (69.0) | 1159 (65.1) | 0.082 |
| Diabetes (%) | 1664 (51.8) | 772 (53.9) | 892 (50.1) | 0.075 |
| Glaucoma (%) | 835 (26.0) | 373 (26.0) | 462 (26.0) | 0.001 |
| Depression (%) | 1174 (36.6) | 550 (38.4) | 624 (35.1) | 0.069 |
| Osteoporosis (%) | 729 (22.7) | 337 (23.5) | 392 (22.0) | 0.035 |
| RA (%) | 2031 (63.2) | 945 (65.9) | 1086 (61.0) | 0.102 |
| Cancer‡ (%) | 693 (21.6) | 313 (21.8) | 380 (21.4) | 0.031 |
| Anemia (%) | 2162 (67.3) | 993 (69.3) | 1169 (65.7) | 0.077 |
| Asthma (%) | 499 (15.5) | 236 (16.5) | 263 (14.8) | 0.046 |
| Hyperlipidemia (%) | 2752 (85.7) | 1248 (87.1) | 1504 (84.5) | 0.073 |
| BPH (%) | 666 (20.7) | 301 (21.0) | 365 (20.5) | 0.012 |
| Hypertension (%) | 2987 (93.0) | 1346 (93.9) | 1641 (92.2) | 0.066 |
| Hypothyroidism (%) | 931 (29.0) | 429 (29.9) | 502 (28.2) | 0.038 |
| CCI Categories (%) |  |  |  | 1.402 |
| ≤ 5 | 1758 (54.7) | 1234 (86.1) | 524 (29.5) |  |
| 6-10 | 1301 (40.5) | 172 (12.0) | 1129 (63.5) |  |
| > 10 | 153 (4.8) | 27 (1.9) | 126 (7.1) |  |
| Predicted mRS Categories (%) |  |  |  | 0.046 |
| ≥ 4 | 1098 (36.1) | 502 (37.3) | 596 (35.1) |  |
| > 4 | 1944 (63.9) | 843 (62.7) | 1101 (64.9) |  |
| Length of Stay (median (IQR)) | 4 (4) | 4 (5) | 4 (4) | 0.021 |
| 90-day Mortality† (%) | 130 (4.0) | 44 (3.1) | 86 (4.8) | 0.091 |

**Legend:** Characteristics of patients stratified by treatment strategy within 14 days after treatment.
†-90 days after time zero (end of initial usage period), also, we note that this is not a baseline characteristic, but an outcome that we have added to describe the unadjusted differences in the groups
‡-Contains prior history of breast, colorectal, prostate, lung, and endometrial cancer.
\*-indicates a percentage that has been obscured for data privacy
IQR, Inter-Quartile Range; CKD, Chronic Kidney Disease; CHF, Congestive Heart Failure; COPD, Chronic Obstructive Pulmonary Disease; CVD, Cardiovascular Disease; MI, Myocardial Infarction; RA, Rheumatoid Arthritis; BPH, Benign Prostatic Hyperplasia; CCI, Charlson Comorbidity Index; mRS, Modified Rankin Score.

### Outcome: Mortality

The standardized 90-day risk of mortality was 53 events per 1000 (95% CI: 49,61) for those who had continued levetiracetam usage and 62 events per 1000 (95% CI: 57,68) for those who did not continue levetiracetam, which resulted in an RD of −9 events per 1000 patients (95% CI: −12,-5). **Figure 2** shows the standardized survival functions and the risk difference.

**Figure 2.**
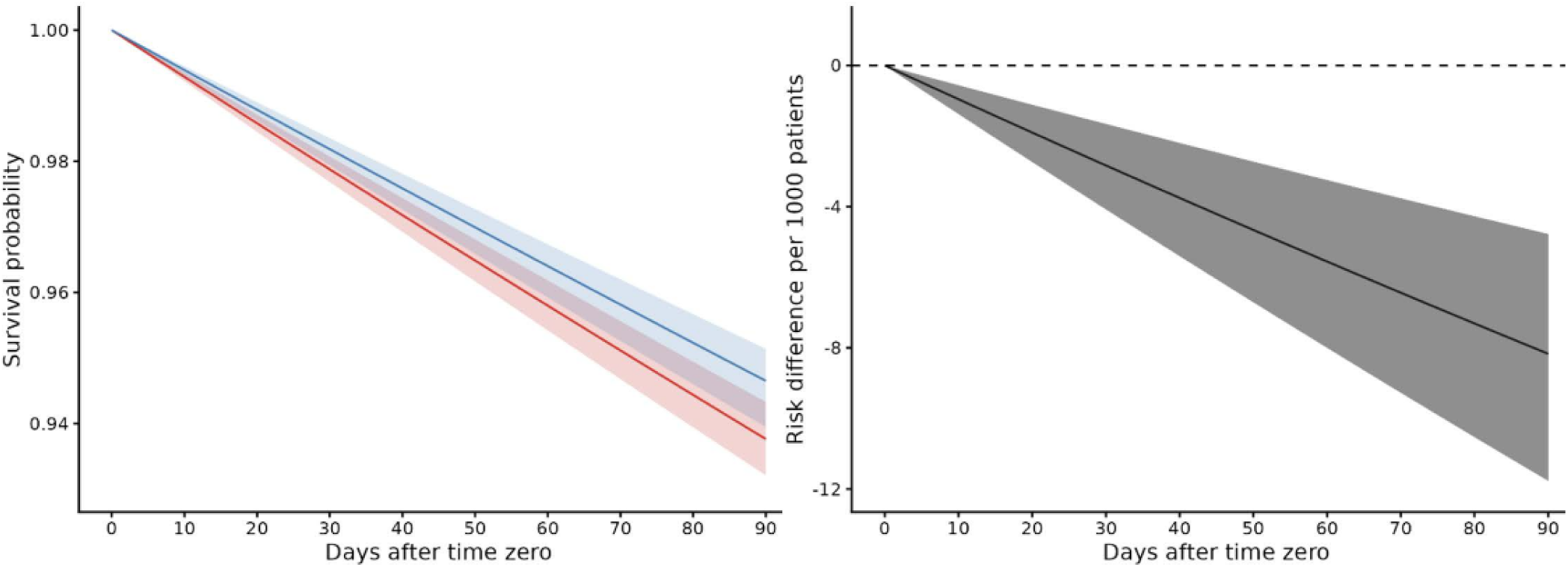
Standardized Survival Curve by Treatment Strategy in Short-Term Initial Usage Sample. **Legend:** Left: Pooled logistic regression survival curves over person-days for all patients. **Blue:** Strategy for continuation of levetiracetam within 14 days after time zero. **Red:** Strategy for no continuation of levetiracetam within 14 days after time zero. Shaded areas: 95% of CIs were constructed using Bootstrap with 500 replications. Right: Risk difference over person-days for all patients. Risk difference between strategies for continuation and no continuation of levetiracetam within 14 days after time zero. Shaded areas: 95% of CIs were constructed using Bootstrap with 500 replications.

**Figure 3.**
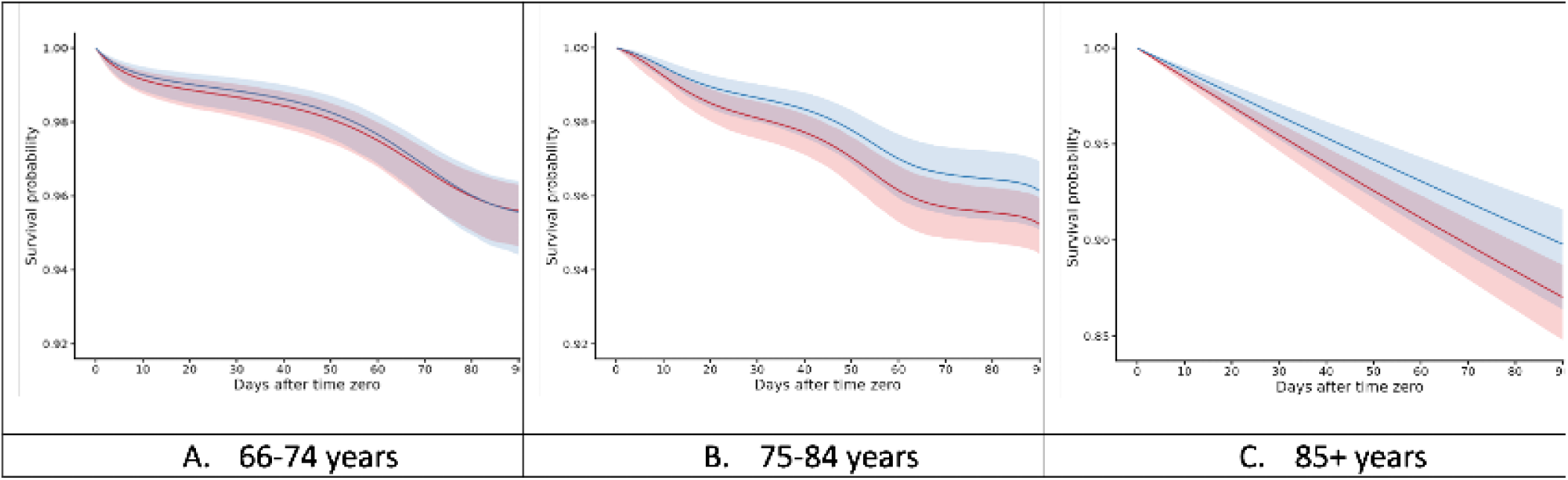
Standardized Survival Curves, Stratified Based on Age in Short-Term Initial Usage Sample. **Legend:** Pooled logistic regression survival curves over person-days for patients of the given strata. **Blue:** Strategy for continuation of levetiracetam within 14 days after time zero. **Red:** Strategy for no continuation of levetiracetam within 14 days after time zero. Shaded areas: 95% of CIs were constructed using Bootstrap with 500 replications.

By age strata, the 90-day RD was 0 events per 1000 (95% CI: −5, +4) for patients aged 66-74, −9 events per 1000 (95% CI: −14, −2) for patients aged 75-84, and −27 events per 1000 (95% CI: −41, −8) for patients aged 85 years or older. We report the standardized 90-day mortality risks stratified by age in **Table 3**.

**Table 3.** Standardized 90-Day Risks of Mortality with Age Stratification in the Short-Term Initial Usage Sample and for Continuers and Discontinuers.

| Age Stratum | Whole Sample |  | Continuers |  | Discontinuers |  |
| --- | --- | --- | --- | --- | --- | --- |
|  | Patients | Mortality | Patients | Mortality | Patients | Mortality |
| *Overall* | 3,212 | 130 | 1779 | 86 | 1,433 | 44 |
| **Age** |  |  |  |  |  |  |
| 66 to 74 years | 860 | 40 | 733 | 32 | 127 | <11 |
| 75 to 84 years | 776 | 36 | 683 | 22 | 93 | 14 |
| 85+ | 405 | 54 | 338 | 32 | 67 | 22 |

| Age Stratum | Continuers |  | Discontinuers |  | Risk Difference |  |
| --- | --- | --- | --- | --- | --- | --- |
|  | Estimate | 95% CI | Estimate | 95% CI | Estimate | 95% CI |
| *Overall* | 53 | (49, 61) | 62 | (57, 68) | -9 | (-12, -5) |
| **Age** |  |  |  |  |  |  |
| 66 to 74 years | 44 | (36, 56) | 44 | (37, 54) | 0 | (-5, 4) |
| 75 to 84 years | 39 | (31, 50) | 48 | (40, 56) | -9 | (-14, -3) |
| 85+ | 102 | (84, 133) | 130 | (113, 152) | -28 | (-39, -3) |

**Legend:** Standardized 90-day risk of mortality (events per 1000 individuals) along with age strata for continuers and discontinuers. CI, confidence intervals.

The risk reduction seems to be concentrated in patients above 75 years of age, particularly over age 85. The analysis in the long-term initial usage sample yielded null results but required a slightly different formulation due to the small sample size (“Additional Analyses” in the Supplement).

## DISCUSSION

In this large, nationally representative emulated target trial of older adults, we examined short-term levetiracetam use after an AIS. After standardization and adjustment for artificial censoring, our results suggest that short–term continuation of levetiracetam among older post–stroke patients may be associated with lower 90–day mortality, particularly in the oldest age groups, while offering no clear survival benefit among younger patients (66-74 years).

In patients with epilepsy, ongoing ASM treatment that reduces generalized tonic-clonic seizure frequency is associated with a lower risk of sudden unexpected death in epilepsy (SUDEP) and epilepsy–related premature death. In contrast, the absence of ASM treatment and poor seizure control increase this risk.^31,32^ In real-world practice, levetiracetam is commonly prescribed during or shortly after AIS and frequently continued beyond discharge.^6^ Our findings suggest that, at least with respect to short-term survival, this continuation strategy does confer measurable benefit. Given the known neuropsychiatric and cognitive adverse effects of levetiracetam, particularly in older adults, the presence of a mortality benefit raises essential questions about the clinical value of routine continuation in this population.^33^ Mortality is a robust and reliably captured outcome in claims-based data and reflects the clinical justification for ongoing ASM (prevention of life-threatening seizures and their downstream consequences).^34,35^

This study is an extension of prior work from our group demonstrating higher short-term mortality associated with initiating epilepsy-specific ASMs within seven days of AIS.^11^ This study provides additional evidence to the existing literature; specifically, it informs patients and providers about the effects of ASMs that have already been started. This study was conducted with additional years of data (up to 2021), additional covariates (such as CCI), and a more valid measure of baseline stroke severity (using predicted mRS). In a previous study using registry-linked electronic health record data, we demonstrated that initiating epilepsy-specific ASMs within 7 days of AIS was associated with a higher 30-day mortality risk than non-initiation, even after extensive confounding adjustment.^1,11^ Findings in this study, however, strengthen the prevailing belief that early poststroke ASM may be beneficial or protective, and challenge clinical judgment to weigh the benefits against the potential risk associated with empiric treatment strategies.^1,11^

Beyond the specific clinical question, this study highlights the methodological value of emulated target trials for epilepsy and stroke pharmacoepidemiology. Conventional observational analyses of poststroke medication effects are highly vulnerable to immortal time bias, confounding by disease severity, and time-dependent treatment assignment.^36,37^ By explicitly defining a target trial of continued versus discontinued levetiracetam use and emulating it using cloning, artificial censoring, and inverse-probability-of-censoring weighting, we aligned treatment assignment with follow-up initiation.^11–13,18^ We preserved the correct temporal ordering of exposure and outcome. This approach enables causal contrasts in settings where randomized trials are infeasible and demonstrates how rich retrospective data can be leveraged to answer clinically consequential questions with good internal validity.^38,39^

From a clinical perspective, these findings have direct implications for medication reassessment and shared decision-making. The widespread continuation of levetiracetam after stroke appears to reflect treatment persistence and lower risk of Sudden Unexpected Death in Epilepsy (SUDEP) and epilepsy–related premature death.^35,40^ However, given the known potential for mood, cognitive, and behavioral adverse effects, routine continuation of levetiracetam in older adults after AIS warrants reconsideration on a case-by-case basis.^41^ Clinicians should be encouraged to consider age as a criterion for reassessing the indication for ongoing ASM and to engage patients and caregivers in discussions about risks, benefits, and goals of care.^42^ Future studies should examine other potential harms, such as cognitive effects or psychiatric issues, which remain understudied. In addition, outcomes such as fall-related injuries, change in mobility or general decrements in quality of life would provide a more complete picture of the pharmacological implications of continued levetiracetam usage.

## Limitations

Our analysis focused exclusively on levetiracetam to ensure precise exposure definition, limiting generalizability to other ASMs and reducing sample size, particularly in long-term initial-use cohorts.^43,44^ Also, claims data do not capture seizure events, EEG findings, lesion characteristics, or neuropsychiatric side effects, precluding direct assessment of non-fatal seizure prevention, seizure burden, other morbidity, or quality-of-life outcomes.^45^

Our definition of continued outpatient use reflects a per-protocol strategy based on prescription fills rather than confirmed medication ingestion, which may introduce exposure misclassification. This information is missing to some extent in the target trial as well but may be captured in pill counts or other quality measures. Additionally, our study does not account for polypharmacy or concurrent use of other ASMs. However, this affects the characterization of the eligible sample; it is expected to attenuate differential effects between treatment strategies rather than generate spurious mortality benefits.

Although emulated target trials substantially reduce bias, residual confounding by unmeasured factors, such as cortical involvement, stroke lesion size, stroke subtype etiology, EEG findings, or clinician prescribing preferences, cannot be excluded.^5,13,46^ Additionally, the statistical estimation of survival functions over a 90-day horizon is contingent on modeling assumptions, and results may be sensitive to outcome model specification; due to the long observation window and large sample size, survival estimates can be influenced by spline specification and functional form choices.^47,48^ We provide detailed justifications and sensitivity analyses regarding model specification and parsimony in the Supplement.

Our findings may not generalize to younger populations, patients with hemorrhagic stroke, Medicare Advantage beneficiaries, or periods beyond 2008 to 2021.^49^ As mentioned above, although mortality is the most robust and reliably captured outcome in claims data, it is an intentionally coarse endpoint. It does not address other clinically significant dimensions of harm or benefit, including seizure recurrence, cognitive trajectories, mood changes, functional recovery, or patient-reported outcomes like quality of life.

## Conclusion

Continued outpatient levetiracetam use among older adults after acute ischemic stroke may be associated with a reduction in 90-day mortality, at least over the short term. These results support a survival benefit of routine levetiracetam continuation after AIS and underscore the importance of deliberate medication reassessment in poststroke care. More broadly, this study demonstrates how emulated target trials can generate ethically feasible, methodologically rigorous evidence to inform epilepsy and stroke care for populations poorly served by conventional randomized trials.

## SUPPLEMENTAL MATERIALS

Tables S1–S5 Figure S1-S8 Technical Details Statistical Code

## Data Availability

The data supporting this study's findings were collected by the Centers for Medicare & Medicaid Services (CMS) and made available without direct identifiers. All results were aggregated in accordance with CMS Cell Suppression Policies. These data are available under license and subject to restrictions. Medicare data are available through CMS with their permission.

